# Disordered balance of T cell subsets in arterial tertiary lymphoid organs in immunoglobulin G4-related vascular disease

**DOI:** 10.1101/2023.03.28.23287890

**Authors:** Satomi Kasashima, Atsuhiro Kawashima, Nozomu Kurose, Satoru Ozaki, Fuminori Kasashima, Yasushi Matsumoto, Hirofumi Takemura, Hiroko Ikeda, Ken-ichi Harada

## Abstract

**Background:** Arterial tertiary lymphoid organs (ATLOs) are ectopic lymphoid structures that control local arterial immune responses and are characterized by germinal centers (GCs). T follicular helper cells (Tfh), resident in GCs, regulate immunoglobulin production and GC development. They consist of Tfh1, Tfh2, and Tfh17 subsets. T follicular regulatory (Tfr) cells possess suppressive functions as Tregs and migrate into GCs as Tfh cells.

Immunoglobulin G4 (IgG4)-related diseases are systemic inflammatory fibrous lesions with elevated serum IgG4 and infiltration of IgG4-positive plasmacytes. These manifest in vascular lesions as frequently formed aneurysms with prominent adventitia thickening (IgG4-related abdominal aortic aneurysm; IgG4-AAA). IgG4-AAAs contain several ATLOs. However, the association of IgG4-AAA pathogenesis and ATLOs or ATLOs and immune cell subsets is poorly understood.

**Methods:** We performed whole-slide immunohistochemical image analysis in surgical specimens of IgG4-AAA (n = 21), non-IgG4-related inflammatory AAA (non-IgG4-IAAA, n = 17), atherosclerotic AAA (aAAA, n = 10), and Takayasu aortitis (TKA, n = 5). We examined the shape of ATLOs, the T cell subsets within and outside ATLOs, and the histological activity of IgG4-relataed disease.

**Results:** IgG4-AAA was characterized by numerous, large, irregular-shaped ATLOs, and higher numbers of Tfr and Tfh2 cells than Tfh1 cells were present compared to patients with other vascular diseases. The morphological abnormalities (in number, area, and form) of ATLOs in IgG4-AAA and the increased number of Tfr cells are closely related to the activity of IgG4-related disease, including the number of IgG4-positive plasmacytes and the degree of adventitial thickness. All T cell subsets (Treg, Th1, Th2, and Th17) were more enriched within ATLOs than outside ATLOs. In particular, an increase in Tfr cells in IgG4-AAA was associated with ATLO formation. Increased Tfh17 cells were found in TKA, and aAAA and non-IgG4-IAAA were characterized by increased Tfh1 cells.

**Conclusions:** In the classification of vascular lesions, considering the imbalance in T cell subsets, IgG4-AAA should be positioned as adventitial vasculitis with predominant Tfr and Tfh2 cells, accompanied by the abnormal appearance of ATLOs.

**Clinical Perspective:** *What is New?:*

- This prospective study uses a new histopathological method for whole slide analysis. We characterized IgG4-related vascular disease according to imbalances in T cell subsets related to morphological changes of arterial/aortic tertiary lymphoid organs.
- Compared to Takayasu arteritis and atherosclerotic changes, imbalances in T cell subsets—according to the predominance of follicular helper type 2 T cells and follicular regulatory T cells in IgG4-related vascular disease—suggested that IgG4-related vascular disease should be defined as adventitial vasculitis.

*What are the clinical implications?:*

- This relatively straightforward method for diagnosing vascular diseases based on imbalances in specific T cell subsets and arterial/aortic tertiary lymphoid organ changes can inform the pathological diagnosis of vasculitis, such as IgG4-related vascular disease and Takayasu arteritis.
- The hallmark of an imbalance in T cell subsets can enable serological diagnosis of vasculitis and use of circulating T cells to monitor disease activity.

## Introduction

Elevated serum IgG4 levels and chronic fibro-inflammation in affected systemic organs, including the salivary glands, pancreas, and retroperitoneum, characterize immunoglobulin G4-related disease (IgG4-RD).^1-3^ Vascular lesions such as inflammatory abdominal aortic aneurysms (IAAA), a type of aneurysm characterized by marked adventitial fibrous thickening that accounts for 5%–10% of all resected abdominal aortic aneurysms, commonly occur in IgG4-RD.^4,5^ About 50% of IAAA cases are represented by IgG4-related abdominal aortic aneurysms (IgG4-AAA).^4,5^

Tertiary lymphoid organs (TLOs) are ectopic lymphoid structures with morphologic features similar to secondary lymphoid organs that allow homing of naïve cells in the T cell area, the interface between the T- and B-cell zones, and the germinal center (GC) areas that present follicular dendritic cells (FDCs). ^6.7^ TLOs are major control sites enabling immune surveillance and innate immune responses. ^6-8^

Arterial/aortic tertiary lymphoid organs (ATLOs) are TLOs in the media and/or the adventitia of the aorta and artery. Resident macrophages, T cells, B cells, mast cells, and dendritic cells in the adventitia have different distributions inside and outside ATLOs.^9,10^ ATLOs have been described in autoimmunity, microbial infection, vasculitis, and atherosclerosis.^11-13^ ATLOs may serve essential functions in IgG4-AAA because the number of ATLOs of IgG4-AAA is characteristically increased compared to atherosclerotic AAA (aAAA). ^4,5,14^

Regarding TLOs/ATLOs, follicular helper T (Tfh) cells, subsets of Th cells expressing CXCR5, BCL-6, and IL-21, are primarily located in GCs.^15^ Tfh cells control the formation of GCs and drive the maturation of long-lived, high-affinity B cells into memory B cells and plasmacytes to produce IgG.^7^ Corresponding to Th subsets (type1 Th, Th1; type 2 Th, Th2; and type 17 Th, Th17), Tfh cells also consist of at least three subsets (type 1 Tfh, Tfh1; type 2 Tfh, Tfh2; and type 17 Tfh, Tfh17). ^15,16^ Follicular regulatory T cells (Tfr) are a subset of forkhead box P3 (Foxp3)+ regulatory T cells (Tregs) that adopt features of Tfh to enable their migration into GC while maintaining their suppressive function as Tregs for immune cells.^17,18^ The ratio of Tfh and Tfr cells and the balance of Th1, Th2, and Th17 cells have attracted attention in the pathogenesis and progression of several immune diseases.^19,20^

In IgG4-RD, the inflammatory and fibrotic processes are controlled by cytokines secreted mainly by Th2 cells and Tregs. ^21,22^ Additionally, a disrupted balance between Th1 and Th2 cells and increased Th2 and Th17 cell counts may contribute to the pathogenesis of IgG4-RD.^23-27^ Furthermore, the essential role of Tfh cells in IgG4-RD has attracted research attention. ^26,28,29^ Akiyama et al. reported that activated peripheral circulating Tfh2 cell counts are high in IgG4-RD. ^23^ This change was positively correlated with the IgG4-RD responder index, the number of affected organs along with the disease state, and circulating Tfh1 cells, whereas Tfh17 cells did not affect serum IgG4 levels. ^23^ Recent reports on the disordered balance of Tfh cells and Th cells outside the GC showed that it might be closely related to the morphological irregularity of TLOs of IgG4-RD in the salivary gland. ^30,31^

In vascular disease, the association of T cell subsets in ATLOs with the pathogenesis and progression of disease has not been precisely examined. This retrospective study histologically examined Tfh subsets, including Tfh1, Tfh2, and Tfh17 cells, and Tregs using whole-slide analysis in IgG4-AAA compared to aAAA, TKA, non-IgG4-IAAA, and normal aorta.

## Methods

### Case selection

We selected 38 cases of IAAA diagnosed by histologic dense adventitia with severe sclerosing inflammation and fibrosis (>2 mm thick) among patients undergoing surgery for aneurysm at Kanazawa University Hospital and Kanazawa Medical Center (Kanazawa, Japan) between January 2006 and December 2016. We included 21 cases of IgG4-AAAs based on the comprehensive histologic diagnostic criteria for IgG4-related diseases as follows^2^: (1) dense lymphoplasmacytic inflammatory infiltrates with increased numbers of IgG4-positive plasma cells (>50/high-power fields [HPFs; 10 eyepiece, 40 object lenses] and IgG4/IgG ratio of >50%) (Figure 1A–C), (2) storiform pattern of fibrosis, and (3) obliterative vasculitis. The 17 remaining IAAAs were classified into non-IgG4-related IAAAs (non-IgG4-IAAA). No significant differentiation in the radiological findings was observed between IgG4-AAAs and non-IgG4-IAAAs. Ten aAAA cases were selected sequentially between January 2006 and December 2006. The patients with Takayasu arteritis (TKA) were diagnosed according to the American College of Rheumatology 1990 criteria.^32^ Five patients with TKA could be examined histopathologically and were included in this study (Figure 1D–F). aAAA was defined as aneurysms with severe atherosclerosis in the intima and media, thin adventitia of <1 mm, and minimal inflammatory cell infiltration (Figure 1G–1I). Ten normal control aortas (no or slight atherosclerotic abdominal aorta without dilatation) were obtained from consecutive autopsies performed at Kanazawa Medical Center between January and August 2012.

**Figure 1.**
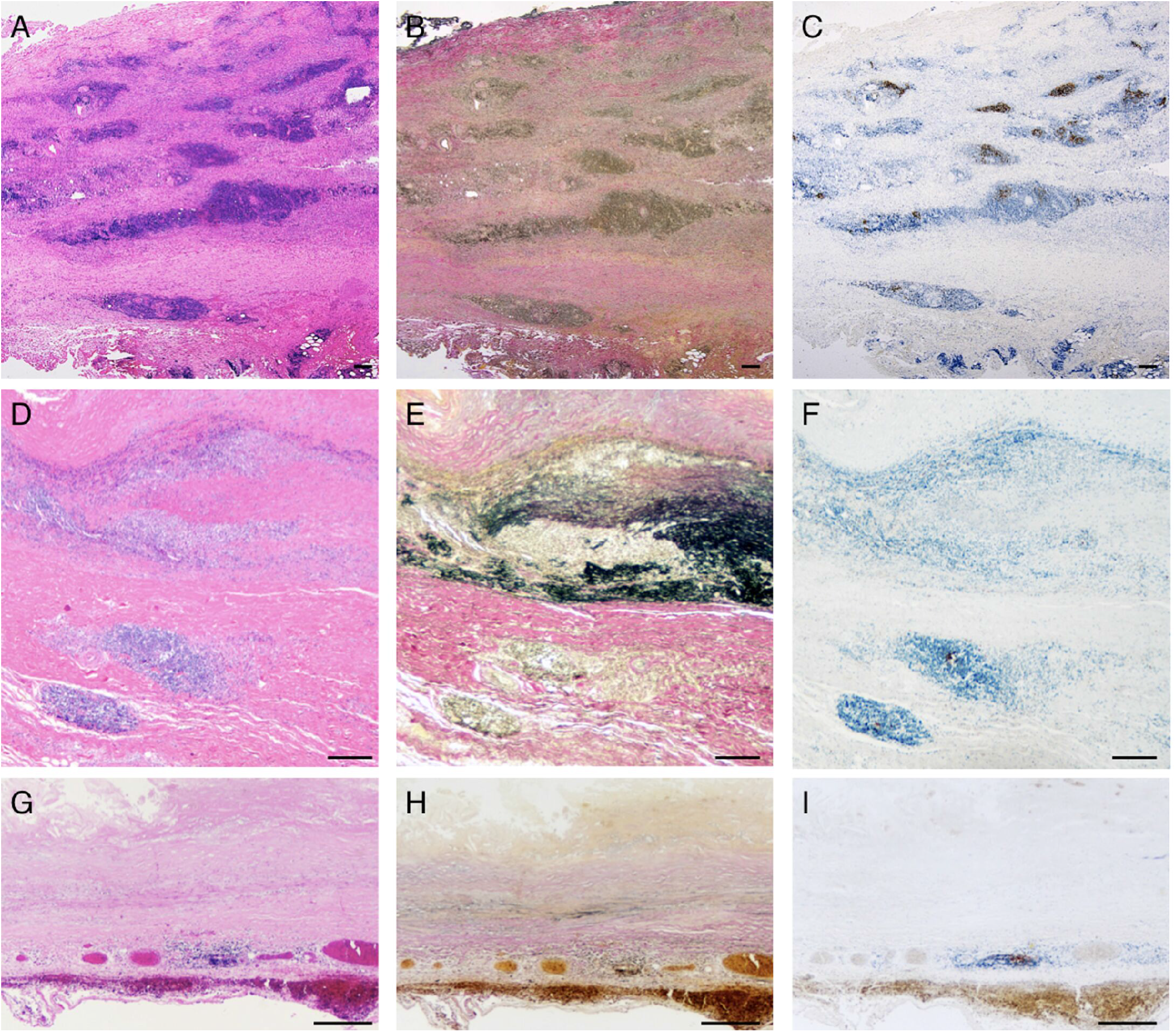
IgG4-related inflammatory abdominal aortic aneurysm (A–C), Takayasu arteritis (D–F), and atherosclerotic abdominal aortic aneurysm (G–I) histopathological features. Characteristic massive fibro-inflammation with lymphoplasmacytic aggregation and thickening adventitia in IgG4-related inflammatory abdominal aortic aneurysm (A, B), degenerative elastic fibers of the media in Takayasu arteritis (D, E), and fibrous intimal thickening and thin media seen in atherosclerotic abdominal aortic aneurysm (G, H). CD21 immunohistochemistory shows follicular dendritic cells distributed across the germinal centers as arterial/aortic tertiary lymphoid organs; large and irregular-shaped germinal centers seen in IgG4-related inflammatory abdominal aortic aneurysm (C); few, small, and oval-to round-shaped germinal centers seen in Takayasu arteritis (F) and atherosclerotic abdominal aortic aneurysm (I). (A, D, and G: Hematoxylin-Eosin staining; B, E, and H: Elastica van Gieson stain; C, F, and I: CD21 immunohistochemistory; A–C: original magnification ×12.5; D–I: original magnification ×20; A–I: scale bar = 500 µm).

In the non-IgG4-IAAA, TKA, aAAA, and control aorta groups, IgG4-positive plasma cells were scarce, and serum IgG4 was within the normal range. Patients who received steroids within 6 months of surgery or biopsy were excluded from all groups. No patient had a ruptured aneurysm or a medical history of another vasculitis (such as giant cell aortitis and antineutrophil cytoplasmic antibody-related vasculitis). The clinicopathological features of patients with IgG4-AAA, non-IgG4-IAAA, TKA, aAAA, and control aorta are summarized in Table 1. Some of these cases were included in our previous reports.^14^

**Table 1.** Clinicopathological features of aAAA, non-IgG4-IAAA, IgG4-AAA, and Takayasu disease.

|  | **aAAA**<br>**(10 cases)** | **non-IgG4-IAAA**<br>**(17 cases)** | **IgG4-AAA**<br>**(21 cases)** | **Takayasu disease**<br>**(5 cases)** |
| --- | --- | --- | --- | --- |
| Age (years) | 68.3 (56–72) | 70.2 (56–81) | 70.5 (70)63–79 | 32.4 (34)22–54 |
| Gender (male/female) | 5/5 | 8/3 | 20/1 | 0/5 |
| Aneurysmal diameter (mm) | 52.1 (34–75) | 49.2 (30–100) | 53.5 (23–87) | 30 (20–45) |
| Low grade fever | 0 | 5 (29.4%) | 10 (47.6%) | 2 (40%) |
| Abdominal or lumbar pain | 1 (10%) | 5 (29.4%) | 15 (71.4%) | 0 |
| C-reactive protein (mg/dL) | 0.41 (0.1–1.1) | 0.69 (0.21–8.04) | 1.44 (0.01–5.22) | 0.78 (0.1–1.51) |
| White blood cell ($\times 10^3/\mu\text{L}$) | 3.4 (3.2–6.7) | 7.9 (3.4–19.5) | 7.1 (4.7–9.5) | 8.9 (6.8–20.2) |
| Serum IgG4 (mg/dL) | 34.5 (12–59) | 24.3 (15.0–58.9) | 91.5 (33.9–559.0) | 23.2 (0–46.9) |
| Serum IgG4/IgG (%) | 2.3 (2.1–2.9) | 1.9 (1.1–6.4) | 8.1 (2.4–29.7) | 2.8 (2.1–3.1) |
| Adventitial thickness (mm) | 0.4 (0.1–1.3) | 3.3 (2.2–6.0) | 4.7 (0.6–14.0) | 1.3 (0.3–1.8) |
| Histological IgG4 (/HPF) | 6.3 (6.1–43.5) | 17.1 (0–41.5) | 83.3 (42.3–175.7) | 12.1 (8.1–22.4) |
| Histological IgG4/IgG (%) | 7.9 (0–38.7) | 26.3 (0–40.9) | 80.2 (58.3–98.7) | 11.4 (3.5–18.4) |

aAAA, atherosclerotic abdominal aortic aneurysm; non-IgG4-IAAA, non-IgG4-IAAA, non-IgG4-related inflammatory abdominal aortic aneurysm
IgG4-AAA, IgG4-related inflammatory abdominal aortic aneurysm; HPF, high power field
data are present as median (range). Three cases of Takayasu disease could measure aneurysmal diameter, serum IgG4, and serum IgG4/IgG.

### Pathological measurements

#### Immunohistochemistry

Immunohistochemistry was performed with a fully automated immunostainer (Roche Diagnostics, Tokyo, Japan; VENTANA BenchMark ULTRA) using antibodies against IgG4 (Nichirei Bioscience, Tokyo, Japan; clone HP6025, ×2), CD21 (Leica Biosystems, Wetzlar, Germany; clone 2G9, ×100) (Figures 2A, 2D), Foxp3 (Abcam; clone236A/E7, ×100) (Figures 2B and 2C), Tbet (Abcam, clone EPR9301, ×200) (Figure 2E), GATA3 (Abcam, clone EPR16651, ×400) (Figure 2F), RORγ (Abcam, clone EPR20006, ×200) (Figure 2G), and BCL-6 (Abcam, Cambridge, UK; clone EPR11410-43, ×1500). CD21 is a marker of FDCs, whereas BCL-6 is a marker of lymphocyte presence in GCs.^8,33^

**Figure 2.**
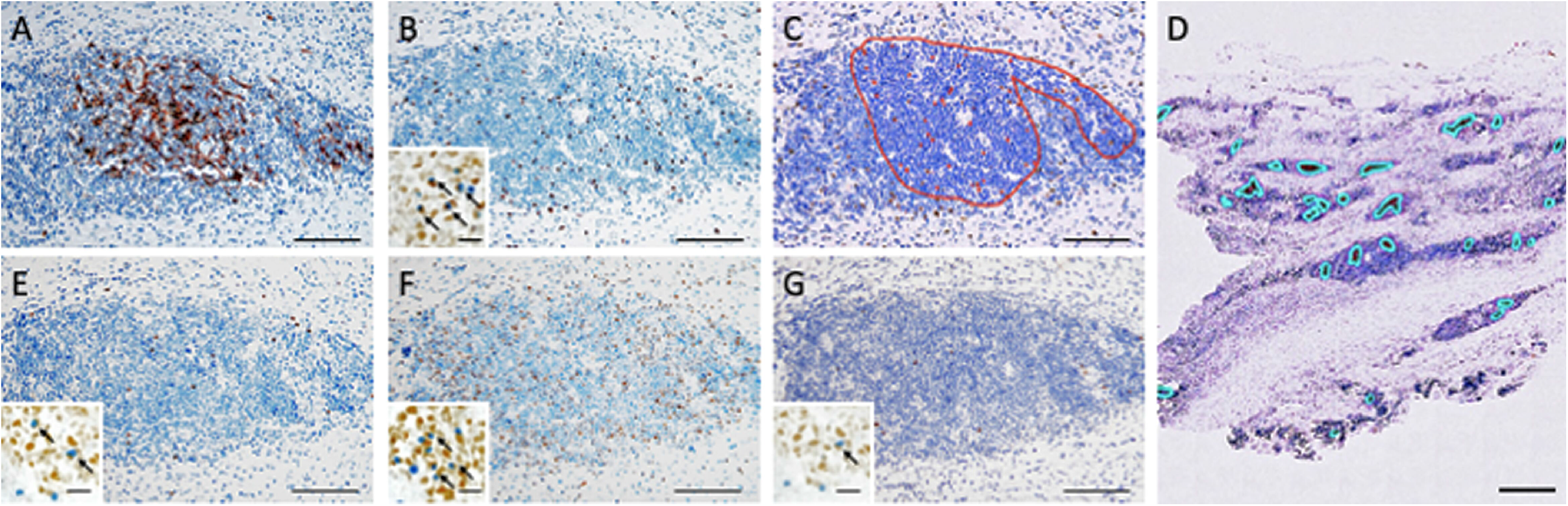
(A–D) QuPath’s methods were used to perform regional annotation using CD21 immunohistochemical-stained slides. The red annotation line is the germinal center’s boundary line and is determined by using CD21 immunohistochemistory (A) and transferred to Foxp3 immunohistochemistory (B). (C) Using QuPath’s cell detection, with the inside red line as the germinal center, Foxp3 immunopositive cells were selected as red-marked cells to calculate the follicular regulatory T-cell number. (D) The Foxp3 immunohistochemistory whole-slide image analysis of IgG4-related inflammatory abdominal aortic aneurysm shows red annotation lines, which are the germinal centers’ boundary lines. Follicular regulatory T cells are calculated in each germinal center, and interfollicular regulatory T cells are calculated outside of germinal centers. T-bet (E), GATA3 (F), and ROR-γ (G) indicate type 1 helper T-cells, type 2 helper T cells, and type 17 helper T-cells, respectively. QuPath’s analyses were performed in the same manner as that for regulatory T cells using Foxp3 immunohistochemistory (A–C and E–G: original magnification ×400, scale bar = 100 µm and D: original magnification ×10, scale bar = 1000 µm).

#### Image data acquisition

All histopathological glass slides were scanned and converted into virtual slides using a whole-slide scanner at ×40 magnification with a resolution of 0.25 µm/pixels (Nano Zoomer, Hamamatsu Photonics, Hamamatsu, Japan).

#### Software

Whole-slide image analysis was performed using QuPath’s interface to the OpenSlide library (https://qupath.github.io). ^34,35^

#### ATLOs analysis

Aggregations of FDCs indicated by CD21 immunopositivity in the media and adventitia were defined as ATLOs (Figures 1C, 1F, and 1I). Using CD21- and BCL-6-immunostained slides, all ATLO regions on each slide were annotated by an experienced pathologist (K.S.) (Figure 2D). A batch script was subsequently applied to automatically identify the area and circumference of each ATLO and the number of ATLOs in the whole-slide images. The circularity of each ATLO was defined as 4πS/L^2^, wherein S and L are the area and circumference, respectively. The circularity of a circle is 1, and decreased circularity of the shape refers to the shape’s irregularity. The area of the ATLOs was calculated as the ratio of the ATLO area to the whole aortic wall area.

#### Analysis of T cell subsets

Foxp3, Tbet, GATA3, and RORγ expression was analyzed using QuPath’s ‘simple tissue detection’ and ‘fast cell counts’ commands. The number of positive cells and the area were used to calculate the average number of positive cells/mm^2^.

#### Definition of T cell subsets

In contrast to Tfr and Tfh cells, T cells outside ATLOs in the adventitia were defined as interfollicular T (ifT) cells. ATLO annotations using CD21-immunostained slides were reconstructed to Foxp3-, Tbet-, GATA3-, and RORγ-stained slides, which were continuous section slides. QuPath commands were used to calculate the numbers of Foxp3-, GATA3-, Tbet-, and RORγ-immunopositive cells within the ATLO areas in the whole-slide images. Foxp3-immunopositive cells distributed within ATLOs were defined as Tfr cells, and those outside ATLOs were considered ifTreg cells. Tbet-immunopositive cells distributed within ATLOs were defined as Tfh1 cells, and those outside ATLOs were considered ifTh1 cells. GATA3-immunopositive cells distributed within ATLOs were defined as Tfh2 cells, and those outside ATLOs were considered ifTh2 cells. RORγ-immunopositive cells distributed within ATLOs were defined as Tfh17 cells, and those outside ATLOs were considered ifTh17 cells. Then, Tfr cells were confirmed by double immunostaining for Foxp3 and BCL-6; Tfh1 cells were confirmed by double immunostaining for Tbet and BCL-6; Tfh2 cells were confirmed by double immunostaining for GATA3 and BCL-6; and Tfh17 cells were confirmed by double immunostaining for RORγ and BCL-6. After this, the ratios of Tfr/if Treg, Tfh1/ifTh1, Tfh2/ifTh2, and Tfh17/ifTh17 cells were calculated.

### Statistical analysis

Kruskal–Wallis analysis of variance was used to compare the groups. Multivariate logistic regression analysis was performed to determine factors that had a significant effect in the univariate analysis and calculate the adjusted odds ratio (OR) and 95% confidence interval (CI). Spearman’s correlation coefficients were used to test the associations between continuous variables. All analyses were performed using SPSS v.28 (IBM, Armonk, NY, USA). Statistical significance was indicated by *p* < 0.05.

### Ethical approval and consent to participate

The study was approved by the Human Investigation Review Committee of Kanazawa Medical Center (No. 30-63) and Kanazawa University Hospital (No. 973-1). It conformed to the principles outlined in the Declaration of Helsinki. The patients provided informed consent before the study.

## Results

### ATLOs

ATLOs were rarely observed in the control aorta (10%) but were sometimes seen in aAAA (40%) and TAK cases (60%). However, all IgG4-AAA samples possessed several ATLOs. ATLOs seen in IgG4-AAA comprised a thicker network of FDCs and were large, distorted, numerous, and evenly distributed in the adventitia (Figure 1C). In ATLOs of the IgG4-AAA group, immunoblasts were sparse, the polarization of recognizable light and dark zones was unclear, and they often collapsed and fused. In contrast, ATLOs seen in the other groups were small and nearly round (Figures 1F and 1I).

The median area and median area rate of the ATLOs were significantly higher in the IgG4-AAA group (2.3 mm^2^ and 1.29%, respectively) than the non-IgG4-IAAA (1.1 mm^2^ and 0.30%, respectively), TKA (0.88 mm^2^ and 0.22%, respectively), aAAA (0.31 mm^2^ and 0.53%, respectively), and normal aorta (0.12 mm^2^ and 0.03%, respectively) (Figure 3). The median circularity of the ATLOs in the IgG4-AAA group (0.64) was significantly lower than that in the non-IgG4-IAAA (0.71, *p* = 0.005), TKA (0.95, *p* = 0.001), aAAA (0.93, *p* < 0.001), and normal aorta (0.95, *p* < 0.001) groups. The median number of ATLOs was higher in the IgG4-AAA group (3.7/mm^2^). However, there was no significant difference between each subgroup. The median number, area, and area rate of ATLOs in the aAAA group were nearly intermediate between those of the normal aorta and non-IgG4-IAAA groups.

**Figure 3.**
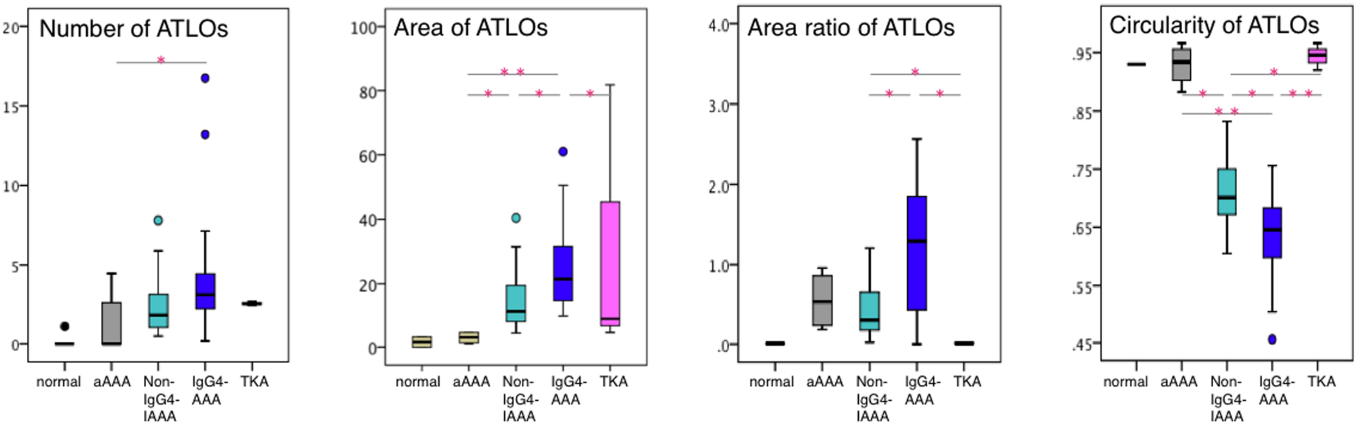
The morphological features of arterial/aortic tertiary lymphoid organs in patients with IgG4-related inflammatory abdominal aortic aneurysm (IgG4-AAA, n = 21), non IgG4-related inflammatory abdominal aortic aneurysm (Non-IgG4-AAA, n = 17), Takayasu arteritis (TKA, n =5), atherosclerotic abdominal aortic aneurysm (aAAA, n = 10), and control aorta (n = 5). Statistically significant differences between groups are indicated at * and **p < 0.01.

### Number of T cells within ATLOs (Tfr, Tfh1, Tfh2, and Tfh17 subsets) in the whole aortic wall

The median number of Tfr cells was significantly higher in the IgG4-AAA group (448.5/mm^2^) than in the non-IgG4-IAAA (211.8/mm^2^, *p* = 0.007), aAAA (1020/mm^2^, *p* = 0.002), and TKA (11.9/mm^2^, *p* < 0.001) groups (Figure 4A). The median number of Tfr cells in the TKA group was significantly lower than that in the IgG4-AAA (*p* = 0.001) and aAAA (*p* = 0.047) groups.

**Figure 4.**
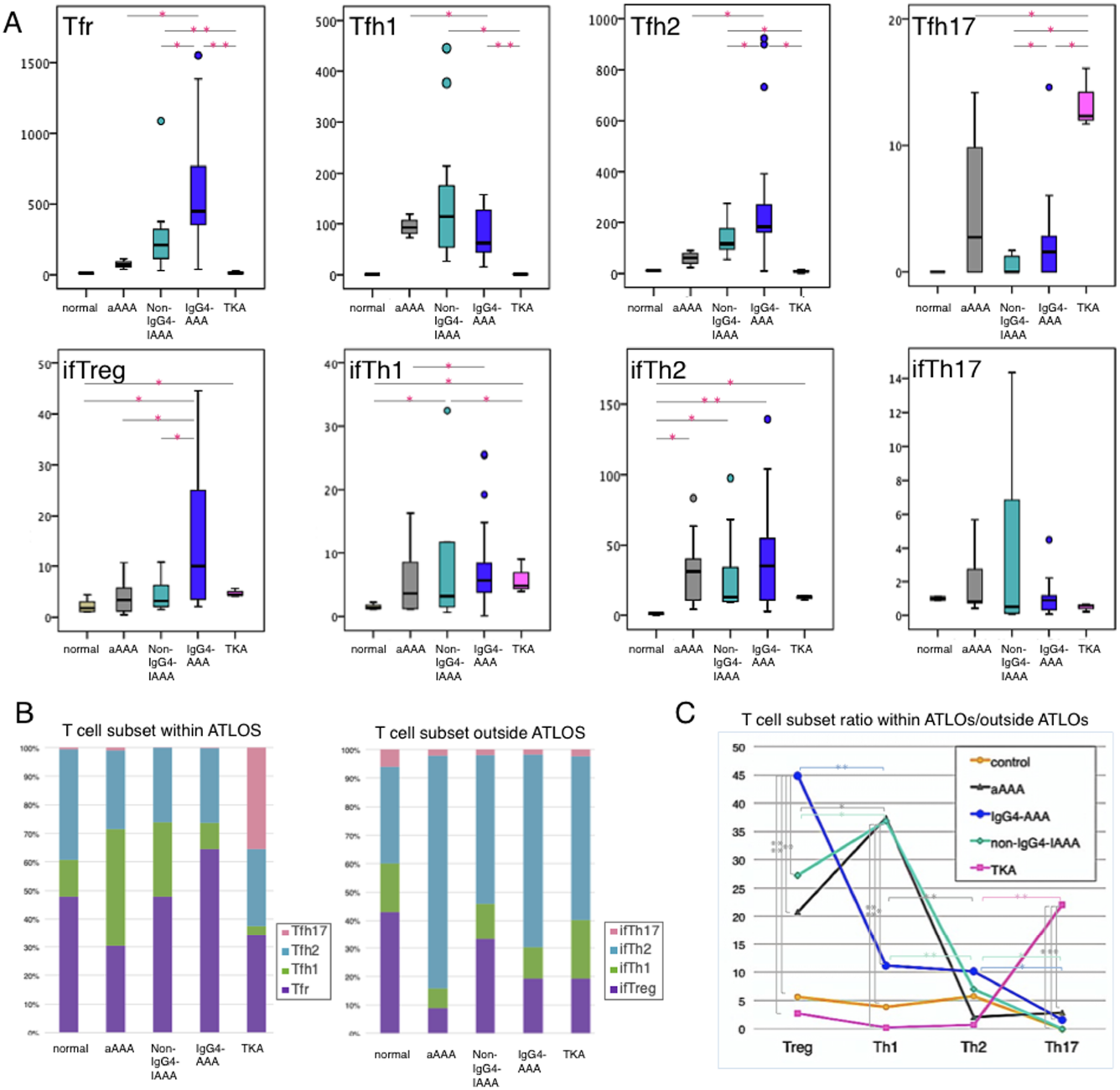
(A) The numbers of T cells subsets in patients with IgG4-related inflammatory abdominal aortic aneurysm (IgG4-AAA, n = 21), non IgG4-related inflammatory abdominal aortic aneurysm (Non-IgG4-AAA, n = 17), Takayasu arteritis (TKA, n = 5), atherosclerotic abdominal aortic aneurysm (aAAA, n = 10), and control aorta (n = 5). (B) The ratio of each T cell subset and (C) the ratio of T cell subsets within ATLOS and outside ATLOs. Tfr, follicular regulatory T cell; Tfh1, type 1 follicular helper T cell; Tfh2, type 2 follicular helper T cell; fTh17, type 17 follicular helper T cell; ifTreg, interfollicular regulatory T cell; ifTh1, type 1 interfollicular helper T cell; ifTh2, type 2 interfollicular helper T cell; ifTh17, type 17; ATLOs, arterial tertiary lymphoid organs. Statistically significant differences between groups are indicated if *p < 0.05 and **p < 0.01.

The median number of Tfh2 cells was also significantly higher in the IgG4-AAA group (182.4/mm^2^) than in the non-IgG4-IAAA (115.9/mm^2^, *p* = 0.031), aAAA (62.0mm^2^, *p* = 0.008), and TKA (9.33/mm^2^, *p* = 0.01) groups. The median number of Tfh2 cells in the TKA group was significantly lower than that in the IgG4-AAA (*p* = 0.004) and aAAA (*p* = 0.047) groups.

The median number of Tfh1 cells was significantly higher in the non-IgG4-IAAA group (114.3/mm^2^), TKA group (1.11/mm^2^, *p* = 0.007), IgG4-AAA (62.9/mm^2^, *p* = 0.056), and aAAA (92.4/mm^2^, *p* = 0.773) groups. The median number of Tfh1 cells in the TKA group was significantly lower than that in the IgG4-AAA (*p* = 0.001) and aAAA (*p* = 0.047) groups.

The median number of Tfh17 cells was significantly higher in the TAK group (12.3/mm^2^) (Figure 1E, 1H, 1K) than in the IgG4-AAA (1.6/mm^2^, *p* = 0.016), non-IgG4-IAAA (0.58/mm^2^, *p* = 0.007), and aAAA (2.23/mm^2^, *p* = 0.047) groups. The median number of Tfh17 cells in the IgG4-AAA group was higher than that in the non-IgG4-IAAA group (*p* = 0.042).

### Number of T cells outside ATLOs (ifTreg, ifTh1, ifTh2, and ifTh17 subsets) in the whole aortic wall

The median numbers of ifTreg, ifTh1, and ifTh2 cells in the whole aortic wall were higher in the IgG4-AAA group (10.0, 5.6, and 35.1/mm^2^, respectively) than in the other groups, whereas the median numbers of ifTh17 cells in all groups were low (less than 1/mm^2^) with no significant differences

### The ratio of T cells within to outside ATLOs (Tf/if T)

In all groups, the numbers of intrafollicular T cell subsets were greater than the corresponding number of extrafollicular T cell subsets; the density of each T cell subset increased within ATLOs (Figure 4B, 4C). However, the concentration of T cell subsets within ATLOs differed in each group of vascular lesions, whereas the ratios of intrafollicular/interfollicular T cells of all T cell subsets were equally very low in the control aorta.

The median Tfr/ifTreg ratio was significantly and characteristically higher in the IgG4-AAA group (44.9) than in the other groups. The median Tfh1/ifTh1 ratio was similarly high in the aAAA (37.4) and non-IgG4-IAAA (36.9) and was significantly higher than that in the IgG-AAA group (11.2; *p* = 0.042, *p* = 0.034, respectively). The median Tfh2/ifTh2 ratio was slightly higher in the IgG4-AAA group. However, there were no significant differences in the other groups. The median Tf17/ifTh17 ratio was significantly higher in the TKA group (22.0) than in the other groups. The median Tf17/ifTh17 ratios in the remaining four groups were low (controls, 0; aAAA, 2.8; non-IgG4-IAAA, 0.1; IgG4-AAA, 1.6).

In the control aorta, the median ratios of intrafollicular/interfollicular T cells were low for all T cell subsets (Tfr/ifTreg ratio, 5.6; Tfh1/ifTh1, ratio 3.8; Tfh2/ifTh2, ratio 5.7; Tfh17/ifTh17 ratio, 0). In the TKA group, the median ratios of Tfr/ifTreg, Tfh1/ifTh1, and Tfh2/ifTh2 (2.7, 0.2, and 0.7, respectively) cells were the lowest among all groups. The median ratio of intrafollicular/interfollicular T cell subsets was similar between the aAAA and non-IgG4-IAAA groups, and there were no significant differences (aAAA vs. non-IgG4-IAAA, Tfr/ifTreg ratio, 20.8 vs. 27.2; Tfh1/ifTh1 ratio, 37.4 vs. 36.9; Tfh2/ifTh2 ratio, 10.2 vs. 7.0; Tfh17/ifTh17 ratio, 2.8 vs. 0.1).

### Correlations

#### Correlations between IgG4-RD pathological parameters and ATLO features

Among all cases, the number of IgG4+ cells significantly correlated with the average area (*R* = 0.541, *p* = 0.001), area ratio (*R* = 0.491, *p* = 0.003), ATLO irregularity (*R* = 0.517, *p* = 0.014), the number of Tfr cells (*R* = 0.446, *p* = 0.008), and the Tfr/ifTfr ratio (*R* = 0.334, *p* = 0.002) (Figure 5A). The IgG4/IgG ratio correlated with the average area (*R* = 0.504, *p* = 0.009), area ratio (*R* = 0.614, *p* = 0.001), ATLO irregularity (*R* = 0.562, *p* = 0.034), number of Tfr cells (*R* = 0.452, *p* = 0.020), Tfr/ifTfr ratio (*R* = 0.443, *p* = 0.021), and Tfh2/ifTfh2 ratio (*R* = 0.443, *p* = 0.002), is reversely correlated with the number of Tfh1 cells (*R* = -0.394, *p* = 0.047). Adventitial thickness also significantly correlated with the average area (*R* = 0.361, *p* = 0.026), ATLO irregularity (*R* = 0.517, *p* = 0.011), number of Tfr cells (*R* = 0.461, *p* = 0.004), number of Tfh2 cells (*R* = 0.461, *p* = 0.042), and Tfr/ifTfr ratio (*R* = 0.389, *p* = 0.021).

**Figure 5.**
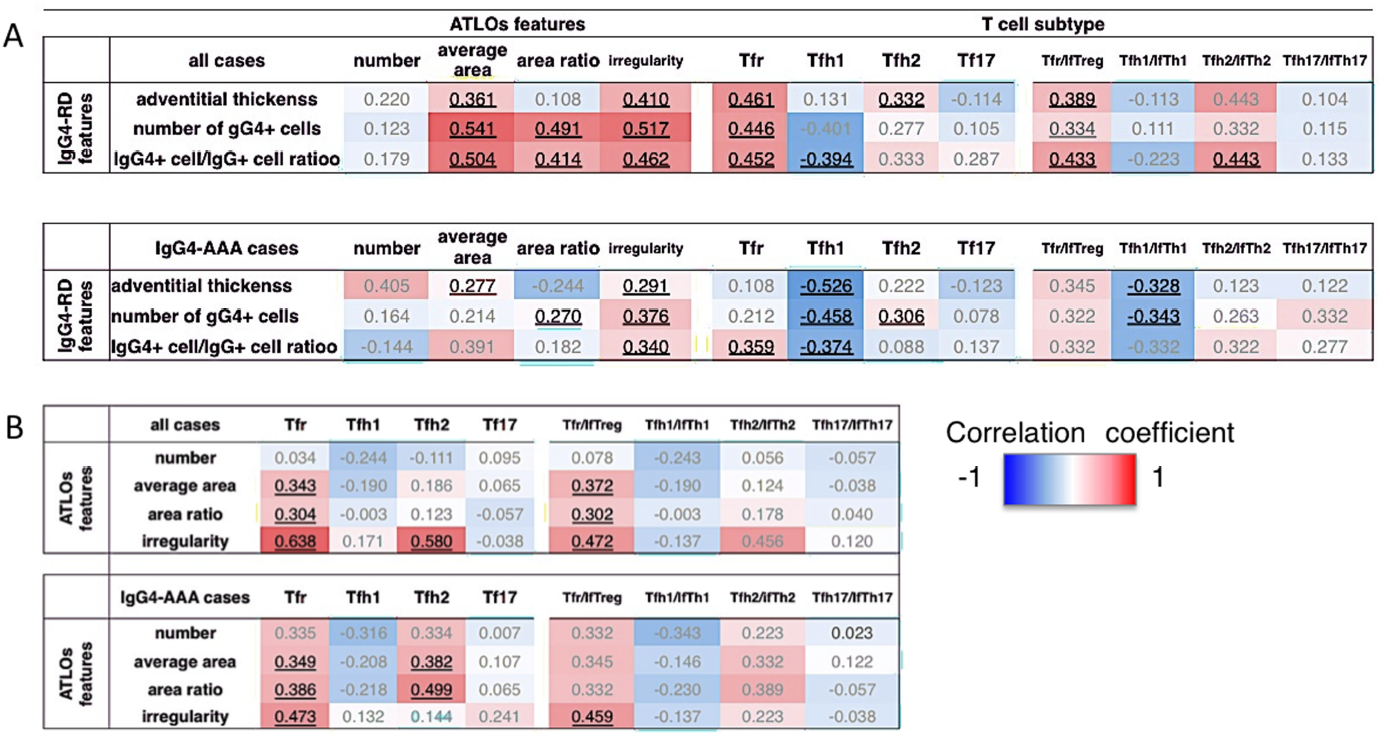
(A) Summary of correlations between IgG4-related disease activity factors and T cell subsets or morphological features of arterial/aortic tertiary lymphoid organs. (B) Summary of correlations between T cell subsets and features of arterial/aortic tertiary lymphoid organs. In each cell, the higher the positive correlation coefficient, the deeper the red color, and the stronger the negative correlation, the deeper the blue. The numbers are correlation coefficients; the bold, dark colored, underlined numbers represent p < 0.05, and the lightly colored numbers are not significant.

Among IgG4-AAA cases, the number of IgG4+ cells significantly correlated with the areal ratio (*R* = 0.270, *p* = 0.048), ATLO irregularity (*R* = 0.338, *p* = 0.030), and the number of Tfh2 cells (*R* = 0.304, *p* = 0.034), is reversely correlated with the number of Tfh1 cells (*R* = -0.458, *p* = 0.037) and Tfh1/ifTfh1 ratio (*R* = -0.343, *p =* 0.003). The IgG4/IgG ratio correlated with ATLO irregularity (*R* = 0.346, *p* = 0.045) is reversely correlated with the number of Tfh1 cells (*R* = -0.458, *p* = 0.040). Adventitial thickness also significantly correlated with the average area (*R* = 0.277, *p* = 0.048) and ATLO irregularity (R = 0.291, *p* = 0.044), is reversely correlated with the number of Tfh1 cells (*R* = -0.526, *p* = 0.036) and the Tfh1/ifTfh1 ratio (*R* = -0.328, *p* = 0.032).

The ATLO irregularity in IgG4-AAA cases was significantly associated with the average area (*R* = 0.442, *p* = 0.021), area ratio (*R* = 0.432, *p* = 0.023), and the number of ATLOs (*R* = 0.331, *p* = 0.034), whereas ATLO irregularity in all cases was not associated with these factors.

#### Correlations between ATLO features and T cell subsets

Among all cases, the average area and area ratio of ATLOs were significantly correlated with the number of Tfr cells (*R* = 0.343, *p* = 0.030; *R* = 0.304, *p* = 0.046, respectively) and the Tfr/ifTreg ratio (*R* = 0.372, *p* = 0.028; *R* = 0.302, *p* = 0.048, respectively) (Figure 5B). ATLO irregularity was significantly correlated with the number of Tfr cells (*R* = 0.638, *p* < 0.001), Tfr/ifTreg ratio (*R* = 0.472, *p* = 0.003), and the number of Tfh2 cells (*R* = 0.580, *p* < 0.001). Among IgG4-AAA cases, the average area (*R* = 0.349, *p* = 0.043), areal ratio (*R* = 0.386, *p* = 0.044), and ATLO irregularity (*R* = 0.473, *p* = 0.022) significantly correlated with the number of Tfr cells (*R* = 0.495, *p* = 0.043). The area ratio was significantly related to the number of Tfh2 cells (*R* = 0.499, *p* = 0.027).

### Multivariable logistic regression

Compared with aAAA, IgG4-AAA was associated with ATLO irregularity, with an adjusted OR of 16.61 (95% CI; 12.34–2.6132, *p* = 0.001), the area ratio of ATLOs, with an adjusted OR of 6.43 (95% CI; 0.345–0.781, *p* = 0.11); the number of Tfr cells, with an adjusted OR of 2.55 (95% CI; 0.894–0.984, *p* = 0.01); and the Tfr/ifTreg ratio, with an adjusted OR of 2.01 (95% CI; 0.789–0.967, *p* = 0.023).

Compared with the other IgG4-AAA cases, IgG4-AAA was significantly associated with ATLO irregularity, with an adjusted OR of 18.95 (95% CI; 231.8–1.28, *p* = 0.006) and the number of Tfr cells, with an adjusted OR of 5.21 (95% CI; 1.99–3.55, *p* = 0.008).

## Discussion

Recently, several reports have indicated that a disrupted balance of T cell subsets contributes to the systemic and vascular manifestations of vascular disease. Th1 and Th17 cytokine responses correlate with TAK activity.^36^ An increase in serum IL-6 characterizes TAK, and IL-6 is a key cytokine involved in the differentiation of Th17 cells and regulates the balance between Th17 and Treg cells. Gao et al. reported that abnormal T cell maturation into Th2-like Tregs and decreased circulating mature Tregs could contribute to TKA pathogenesis.^37^ Our findings demonstrate that the number of Tfh17 cells was higher in TKA than in other vascular groups. The increase in Th17 cells in TAK might be due to the higher number of Tf17 cells.

Flow cytometry-based peripheral blood analysis revealed that uncontrolled inflammation involving Tfh and Tfr cells underlies IgG4-RD etiology.^38^ A disrupted balance between Th1 and Th2 cells and increased Th2 and Th17 cell counts may contribute to IgG4-RD pathogenesis.^15,23-28^ Grados et al. reported that IgG4-RD is characterized by a shift in circulating T cells toward Th2/Tfh2 and Th17/Tfh17 polarization.^39^ However, previous studies of circulating T cells via flow cytometry have not evaluated intrafollicular T cells because Tfr and Tfh cells present in GCs have different phenotypes from circulating Tfr and Tfh cells, which are primarily defined by CXCR5 positivity.^20^

Using whole-slide analysis, this is the first report to describe the T cell subsets in IgG4-AAA, particularly regarding ATLOs. In IgG4-AAA, the number of Tfr, ifTreg, Tfh2, and ifTh2 cells were significantly high; therefore, all Treg and Th2 cells were also more numerous than other groups. Moreover, the Tfr/ifTreg ratio was considerably higher than that of the other groups. The Tfr cell count and the Tfr/ifTreg ratio were closely related to IgG4-AAA activity (adventitia thickness, IgG4+ cell count, and IgG4/IgG ratio). Thus, the increase in Treg cell concentrations within ATLOs was closely associated with the pathogenesis and progression of IgG4-AAA and is a novel pathological hallmark of IgG4-AAA. Regarding T cell subsets, the Tfh2 cell number increase and Tfh1 cell number decrease might assist in the pathological diagnosis of IgG4-AAA. Reliable pathological hallmarks of IgG4-AAA are needed given that diagnosis of IgG4-RD using serum IgG4 levels or tissue IgG4+plasmacytes is challenging. The number of circulating Tfr cells or the Tfr/Treg ratio, not just Treg cells, should be assessed to diagnose or estimate the activity of IgG4-AAA using flow cytometry. Considering that the interaction of Tfr and Tfh cells regulates the production of IgG within the GC, controlling Tfr cells creates opportunities to develop novel therapeutic approaches in immune disease^40,41^. Our findings indicate that Tfr therapy might be a promising therapeutic method for IgG4-AAA in the future.

ATLOs are rarely present in the normal aorta, and their presence suggests that there is a vascular lesion. Previous reports indicated that ATLOs may organize pro- and atherogenic immune responses, and disruption in the balance of immune cell subsets may trigger atherosclerosis. ^42,43^ In this study, the ATLOs in aAAA were sparsely distributed, small, and nearly round, similar to TLOs in chronic sialadenitis. These are thought to be a response to local chronic stimulation. ^43^ Clemet et al. noticed that active TKA with increased aortic 18F-fuluoro-deoxyglucose uptake contained several ATLOs, whereas a clear association between pathogenesis and ATLOs in TKA was not observed.^13^ In contrast, ATLOs in IgG4-AAA were significantly larger, more numerous, and more irregular-shaped, with a thick network of FDCs, compared to other vascular groups. Larger ATLO size in IgG4-AAA was associated with a more irregular shape and a higher number of ATLOs. Additionally, they were fused, or divided and collapsed, not simply enlarged. Moreover, the area, number, and ATLO irregularity in IgG4-AAA significantly correlated with the activity of IgG4-RD, such as the number of IgG4+ plasmacytes and serum IgG4 levels. Therefore, we believe that the morphological deviation of ATLOs in IgG4-AAA might be associated with the active phase of IgG4-AAA,^14^ which could be useful for the pathological diagnosis of IgG4-AAA. Recently, inflammation evaluated by 18F-PET/CT has been noted in IgG4-related vascular diseases.^44^ Based on TKA,^36^ the activity of IgG4-AAA evaluated by FDG uptake observed by PET/CT might reflect the amount or activity of ATLOs in IgG4-AAA.

Tfr cells are more likely to regulate the function and output of TLOs/ATLOs at an established GC. In contrast, ifTreg cells control initial GC formation; hence, the balance between Tfr and ifTreg cells could contribute to GC development and decay by regulating the production of several cytokines, such as IL-10 and IL-21.^38,45^ In IgG4-SS, Tfr/ifTreg imbalance is associated with the invagination of the surrounding T–B-cell border into TLOs, causing TLOs to have irregular shapes.^30,31^ Similarly, in IgG4-AAA, both the number of Tfr cells and the Tfr/if Treg ratio are closely related to the average area, area ratio, and ATLO irregularity. Thus, the Tfr/ifTreg imbalance reflects characteristic ATLOs dysplasia in IgG4-AAA.

The morphological dysplasia of TLOs in IgG4-SS is greater (larger and more irregular ATLOs with more prominent immunoblasts) than the ATLOs in IgG4-AAA.^30,31^ Compared to IgG4-AAA, IgG4-SS cases might be in a more active phase; they often have high serum IgG4 levels and are prone to multiple organ lesions of IgG4-RD. In IgG4-SS, the Tfh2/ifTh2 and Tfh17/ifTh17 ratios are high, and the Tfh17/ifTh17 ratio is significantly related to the number of IgG4 cells in the area, the area size, and TLO irregularity. ^31^ In contrast, IgG4-AAA shows a higher degree of fibrosis and is more closely related to inflammatory features, including elevated serum CRP and IL-6 levels related to local IL-6 production.^14^ This report illustrates that in IgG4-AAA, the number of Tfh17 cells and the Tfh17/ifTh17 ratio were low, and the Th17 cell component was not related to IgG4-AAA activity and ATLO morphology. Therefore, the differences in the morphological abnormalities of ATLOs/TLOs in IgG4-AAA and IgG4-SS may be due to differences in the activity of IgG4-RD and pathogenesis and progression factors.

The number of each T cell subset in aAAA was uncharacteristic. However, the ratios of intrafollicular/interfollicular T cell subsets in aAAA were strikingly different from those of IgG4-AAA, showing a moderate increase in the Tfr/ifTreg ratio, a high Tfh1/ifTh1 ratio, and low ratios of Tfh2/ifTh2 and Tfh17/ifTh17. Interestingly, the ratios of intrafollicular/interfollicular T cell subsets in non-IgG4-IAAA were similar to those of aAAA, while the number of each T cell subset in non-IgG4-IAAA was higher than those of aAAA. Thus, aAAA and non-IgG4-IAAA were similar, and non-IgG4-IAAA might be an accelerated inflammatory state of aAAA.

In conclusion, the increases in T cell subsets inside ATLOs were characteristic for each type of vascular lesion. Tfr and Tfh2 cells were predominant over Tfh1 cells in IgG4-AAA. The increase in Tfr and the decrease in Tfh1 cells represent novel hallmarks of IgG4-AAA that are associated with morphologically abnormal ATLO formation and disease pathogenesis. An increase in Tfh17 cells was an attribute of TKA, and aAAA and non-IgG4-IAAA were characterized by an increase in Tfh1 cells. In the classification of vascular lesions, based on the imbalance in T cell subsets, IgG4-AAA should be positioned as vasculitis in predominant adventitia accompanied by morphological abnormalities of ATLOs.

## Data Availability

Whole-slide image analysis was performed using QuPath's interface to the OpenSlide library

https://qupath.github.io

## Nonstandard abbreviations and acronyms

(IgG4-AAA): immunoglobulin G4-related abdominal aortic aneurysm
(non-IgG4-IAAA): non-immunoglobulin G4-related inflammatory abdominal aortic aneurysm
(ATLOs): arterial/aortic tertiary lymphoid organs
(Tfh): follicular helper T cell
(Th1): type 1 helper T cell
(Th2): type 2 helper T cell
(Th17): type 17 helper T cell
(Tfh1): type 1 follicular helper T cell
(Tfh2): type 2 follicular helper T cell
(Tfh17): type 17 follicular helper T cell
(ifTh1): interfollicular type 1 helper T cell
(ifTh2): interfollicular type 2 helper T cell
(ifTh17): interfollicular type 17 helper T cell
(Tregs): regulatory T cells
(Tfr): follicular regulatory T cells
(ifTreg): interfollicular regulatory T cells

## Acknowledgements

The authors would like to sincerely thank the Master’s students Madoka Igarashi, Miyu Okuda, Ms. Hiromi Kojima, and Ms. Youko Umehara for their technical assistance.

## Declarations

### Author Contributions

SK performed data selection, experiments, data analysis, and manuscript writing. NK, FK, YM, and HI participated in the selection of cases. AK and KH supervised the work and were involved in setting up the experimental design. All authors reviewed the manuscript and agreed with its contents.

